# Elevated Plasma Monounsaturated Fatty Acids and Their Associations with Disease Activity, Adiposity, and Sex in Patients with Rheumatoid Arthritis: A Cross-Sectional Study

**DOI:** 10.64898/2026.01.27.26344951

**Authors:** Shalini N. Swamy, Martha A. Belury, Rachel M. Cole, Kristen Heitman, Steven Pan, Zhibo Yang, Aizhan Karabukayeva, Yang Mao-Draayer, Beatriz Y. Hanaoka

## Abstract

**Background:** Rheumatoid arthritis (RA) is a chronic inflammatory disease characterized by metabolic dysregulation, including altered lipid metabolism. While polyunsaturated fatty acids have been studied, the plasma levels, endogenous synthesis, and relevance of monounsaturated fatty acids (MUFAs) in RA remain unclear. This study examined plasma MUFA levels in RA and their associations with disease activity, adiposity, and intake.

**Methods:** In this cross-sectional study, 59 individuals with rheumatoid arthritis (RA) and 33 non-RA controls frequency-matched on age, sex, and BMI were recruited between 2017 and 2022. Clinical assessments included disease activity (DAS28), body composition, and metabolic parameters. Dietary intake was assessed using a 4-day food journal, and plasma fatty acids were quantified by gas chromatography in 82 participants with available samples. The stearoyl-CoA desaturase-1 (SCD-1) index was used as a proxy for endogenous MUFA synthesis. Associations between MUFAs and clinical variables were evaluated using univariate and multivariable regression (p<0.05).

**Results:** RA participants had higher waist-to-hip ratio, fat mass, fasting triglycerides, and lower physical activity than controls. Plasma palmitoleic and oleic acids and the SCD-1 index were higher in RA, whereas linoleic and arachidonic acids were lower. Saturated and omega-3 fatty acids were similar. Higher oleic and gondoic acids were independently associated with greater disease activity; oleic acid was linked to central adiposity, and palmitoleic acid was higher in women, suggesting sex– and adiposity-specific regulation.

**Conclusions:** Higher plasma MUFAs in RA are associated with disease activity, adiposity, and sex, highlighting altered MUFA metabolism as a feature of RA and a potential target for metabolic intervention.

**Key Messages:** *What is already known on this topic:* Rheumatoid arthritis (RA) involves systemic inflammation and altered lipid metabolism. While polyunsaturated fatty acids have been studied extensively, the plasma levels, endogenous synthesis, and clinical relevance of monounsaturated fatty acids (MUFAs) in RA remain unclear.

*What this study adds:* Patients with RA have higher plasma MUFAs, including oleic and palmitoleic acids, and an elevated SCD-1 index, a marker of endogenous MUFA synthesis. Higher MUFAs are associated with disease activity, central adiposity, and sex-specific patterns, independent of dietary intake.

*How this study might affect research, practice or policy:* Plasma MUFAs could serve as potential biomarkers of RA disease activity and metabolic dysregulation. These findings suggest that altered MUFA metabolism contributes to inflammatory pathways, highlighting a potential target for future research, nutritional interventions, or therapeutic strategies.

## Introduction

Rheumatoid arthritis (RA) is a systemic autoimmune disease characterized by chronic synovial inflammation and associated metabolic dysfunction(1). Its pathogenesis involves a complex interplay of genetic, environmental, and immunological factors that promote aberrant immune activation and persistent inflammation(2). Alterations in plasma fatty acid composition have been linked to immune dysregulation in RA and other diseases(3).

Polyunsaturated fatty acids (PUFAs) are among the most extensively studied fatty acids in RA. N-6 fatty acids, particularly arachidonic acid, serve as precursors for both pro-inflammatory eicosanoids (e.g., prostaglandins and leukotrienes) and anti-inflammatory lipoxins, while n-3 fatty acids give rise primarily to anti-inflammatory and pro-resolving mediators such as resolvins, protectins, and maresins(4) (5). Observational data suggest that a higher dietary n-6 to n-3 PUFA intake ratio may be associated with greater pain in methotrexate-treated early RA patients, despite well-controlled inflammation as indicated by DAS28, CRP, and ESR (6). In contrast, supplementation with n-3-rich fish oil in patients with RA has been reported to reduce inflammation, improve joint function(7), decrease the need for DMARDs and NSAIDs(8), and may slow disease progression(9).

Beyond PUFAs, other fatty acids modulate immune pathways, and may have similar effects in RA. Among monounsaturated fatty acids (MUFAs), oleic acid (18:1n9) has demonstrated anti-inflammatory effects in experimental studies and may exert similar anti-inflammatory effects in humans(10). Oleic acid (18:1n9) and palmitoleic acid (16:1n7) are the most abundant MUFAs in humans and arise from both dietary intake and endogenous metabolism(11).

When dietary intake is limited or metabolic demand is increased, endogenous MUFA synthesis contributes substantially to circulating MUFA levels. This process is regulated by stearoyl-CoA desaturase-1 (SCD-1), the rate-limiting enzyme that converts saturated fatty acids, stearic (18:0) and palmitic (16:0) acids, into oleic and palmitoleic acids (12). The oleic-to-stearic acid ratio (18:1/18:0) is commonly used as a surrogate marker of SCD-1 activity, providing an indirect measure of endogenous MUFA production(13, 14).

SCD-1 is tightly regulated by multiple physiological and environmental factors. Dietary components, including PUFAs and carbohydrates, as well as hormonal signals such as insulin, play a key role in controlling its activity(12). PUFAs generally suppress SCD-1(15), whereas carbohydrates and insulin upregulate it(16), increasing endogenous MUFA synthesis. By modulating MUFA availability, these dietary and hormonal factors may influence systemic inflammation (12).

This study aims to investigate the associations among plasma fatty acids, adiposity, and RA disease activity, with a focus on MUFAs and SCD-1. A better understanding of these relationships may provide new insights into the metabolic regulation of inflammation in RA and identify potential targets for therapeutic intervention.

## Methods

### Study Aim, Design and Setting

This cross-sectional study aimed to investigate plasma monounsaturated fatty acids (MUFAs) in patients with rheumatoid arthritis compared with matched controls and to examine their associations with disease activity, body composition, and endogenous metabolic regulation. The study was conducted at the Division of Rheumatology, University of Alabama at Birmingham; the Division of Rheumatology and Immunology, The Ohio State University; and the Section of Rheumatology, Immunology and Allergy, University of Oklahoma Health Sciences Center. Research participants were enrolled in the study between March 2017 and June 2022. RA participants were recruited from rheumatology outpatient clinics at participating institutions during the period of the study, including while the corresponding author was faculty at the University of Alabama at Birmingham. Standard operating procedures were used across sites for sample processing and storage.

### Participants

This cross-sectional study was approved by the Institutional Review Boards at the participating institutions, and all participants provided written informed consent. Fifty-nine patients diagnosed with rheumatoid arthritis (RA) according to the 2010 ACR/EULAR classification criteria (17) and 33 non-RA controls were enrolled. Non-RA controls were frequency-matched to RA participants on age, sex, and body mass index (BMI) to achieve comparable group distributions.

Participants were excluded if they had other autoimmune inflammatory conditions, chronic viral infections (hepatitis B or C or HIV), type 2 diabetes mellitus, chronic liver or kidney disease, or were pregnant. RA participants were recruited from rheumatology outpatient clinics at participating institutions. Non-RA controls were recruited from community volunteers and were screened to exclude inflammatory arthritis and autoimmune disease. Of these, 82 participants had available plasma fatty acid measurements and were included in the analytic sample. Dietary intake data were available for all participants included in the analyses. For other study variables, missingness was minimal and analyses were conducted using complete-case methods.

### Clinical Assessments

For RA patients, disease duration, and current use of prednisone and disease-modifying antirheumatic drugs (DMARDs) were recorded. Rheumatoid factor (RF) and anti-cyclic citrullinated peptide antibodies (anti-CCP) were measured in the clinical laboratory. Disease activity was assessed using the Disease Activity Score-28 C-reactive protein (DAS28-CRP). Metabolic parameters, including hemoglobin A1c (HbA1c), fasting triglycerides, and insulin sensitivity estimated by the Matsuda index(18) from a 2-hour oral glucose tolerance test (OGTT), were also measured to characterize the metabolic profile of study participants.

### Anthropometric Measurements

Height, weight, waist, and hip circumferences were obtained in accordance with National Health and Nutrition Examination Survey III (NHANES III) anthropometric measurement protocol(19). BMI was calculated as weight (kg) divided by height squared (m²). Waist-to-hip ratio (WHR) was calculated as waist circumference divided by hip circumference. Whole-body dual-energy X-ray absorptiometry (DEXA) was used to assess body composition, from which fat mass index (FMI; fat mass [kg]/height² [m²]) and fat-free mass index (FFMI; fat-free mass [kg]/height² [m²]) were derived.

### Plasma Fatty Acid Analysis

Plasma total lipids were extracted according to the Folch method(20). Fatty acid methyl esters (FAMEs) were prepared using 5% hydrochloric acid in methanol(21). Gas chromatography analysis of FAMEs was conducted using a 30-m Omegawax™ 320 fused silica capillary column (Supelco, Bellefonte, PA). Retention times of samples were compared to fatty acid methyl ester standards (Matreya, LLC, Pleasant Gap, PA, and Nu-Check Prep Inc., Elysian, MN). Fatty acids are reported as a percentage of total identified(22). Stearoyl-CoA desaturase-1 (SCD-1) activity was estimated indirectly using the plasma ratio of oleic-to-stearic acid (18:1n9/18:0), a commonly used proxy for SCD-1 activity (14).

### Physical Activity Assessment

Physical activity was assessed using the short form of the International Physical Activity Questionnaire (IPAQ)(23) to characterize participants’ lifestyle behaviors. Total activity was expressed in MET-minutes per week, calculated according to the IPAQ scoring protocol as the sum of walking, moderate, and vigorous activity.

### Dietary Assessment

Dietary intake data was collected using a 4-day self-reported food journal, which was analyzed with the Nutrition Data System for Research (NDSR) software version 2016 (Nutrition Coordinating Center, University of Minnesota, Minneapolis, MN) to characterize participants’ dietary patterns and quantify intake of fatty acids(24). Diet quality was assessed using the Healthy Eating Index–2015 (HEI-2015), which compares participant dietary intake with the Dietary Guidelines for Americans. The energy adjusted HEI-2015 was calculated using a density-based, energy-adjusted scoring approach (per 1,000 kcal) in accordance with established methodology(25).

### Statistical Analysis

Continuous variables are presented as mean ± standard deviation or median (interquartile range), as appropriate, and categorical variables as counts and percentages. Differences between patients with rheumatoid arthritis (RA) and non-RA controls were assessed using Student’s *t*-test or Wilcoxon rank-sum test for continuous variables and chi-square or Fisher’s exact test for categorical variables.

Associations between plasma monounsaturated fatty acids (MUFAs) and RA disease activity, measured by the Disease Activity Score-28 using C-reactive protein (DAS28-CRP), were evaluated among RA participants using Spearman correlation. Multivariable linear regression models were constructed to assess the independent associations of individual MUFAs with RA disease activity (DAS28-CRP) and, separately, with RA status (RA vs control), adjusting for age, sex, dietary intake of the specific fatty acid, and waist-to-hip ratio.

Regression model assumptions, including linearity, normality of residuals, and homoscedasticity, were examined and met. Sample size was determined by the number of participants enrolled during the recruitment period with available plasma fatty acid measurements and dietary intake data; no a priori power calculation was performed.

Multivariable models were fit using complete-case analysis (participants with missing data for any covariate in a given model were excluded); therefore, sample sizes vary by model. All statistical analyses were conducted using R software (version 4.4.0). A two-sided p-value < 0.05 was considered statistically significant.

### Bias

To reduce confounding, non-RA controls were matched to RA participants on age, sex, and body mass index (BMI), and multivariable regression models further adjusted for age, sex, waist-to-hip ratio, and dietary intake of the corresponding fatty acid. Standardized protocols were used across study sites for anthropometric measurements, dual-energy X-ray absorptiometry (DEXA), and disease activity assessment (DAS28-CRP).

Dietary intake was assessed using a 4-day self-reported food journal, which may be subject to recall and reporting error. Plasma fatty acids were quantified using standardized gas chromatography methods, and laboratory personnel performing fatty acid analyses were blinded to participant RA status.

### Manuscript Preparation

Portions of the manuscript text were drafted and edited with the assistance of a Large Language Model (ChatGPT, OpenAI, San Francisco, CA, USA). All study design, data analysis, interpretation, and final text were reviewed and verified by the authors, who take full responsibility for the content.

## Results

### Participant Characteristics

A total of 59 RA patients and 33 frequency-matched on age, sex, and BMI controls were included in the analysis (Table 1). RA participants had a median age of 56 years (IQR 50–60), were predominantly women (88%), and had a median BMI of 30 kg/m² (IQR 26–35). Matching variables did not differ significantly between groups (all p>0.20). Compared with controls, RA patients had a higher waist-to-hip ratio (0.90 vs 0.86, p = 0.033), higher fat mass index (12.7 vs 10.5 kg/m^2^, p = 0.025), elevated fasting triglycerides (95 vs 71 mg/dL, p = 0.011), and lower physical activity (2,373 vs 4,620 MET-min/week, p = 0.029). HDL cholesterol was numerically lower in RA (p = 0.062), while HbA1c was similar (median 5.4%, p >0.9). These findings reflect a less favorable metabolic profile and lower physical activity levels in RA compared with controls. Among RA participants (Table 2), 76% were seropositive, with a median disease duration of 7 years and low disease activity (median DAS28-CRP 1.77). Prednisone use was low (15%, median dose of 5 mg/day), and 96% were on DMARDs.

**Table 1.**
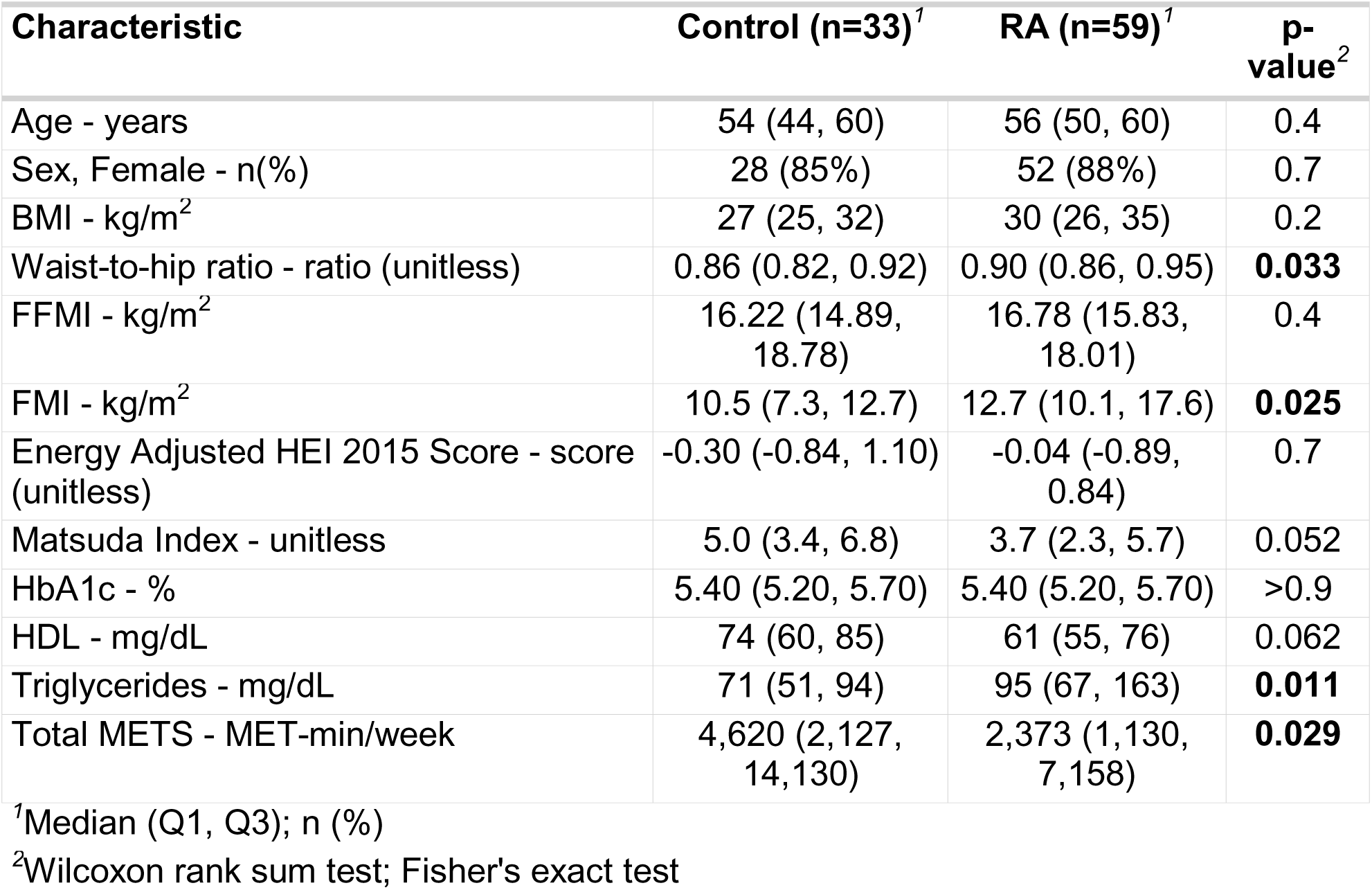
Demographic, Clinical, and Dietary Characteristics of Study Participants by RA status (n=92)

**Table 2.**
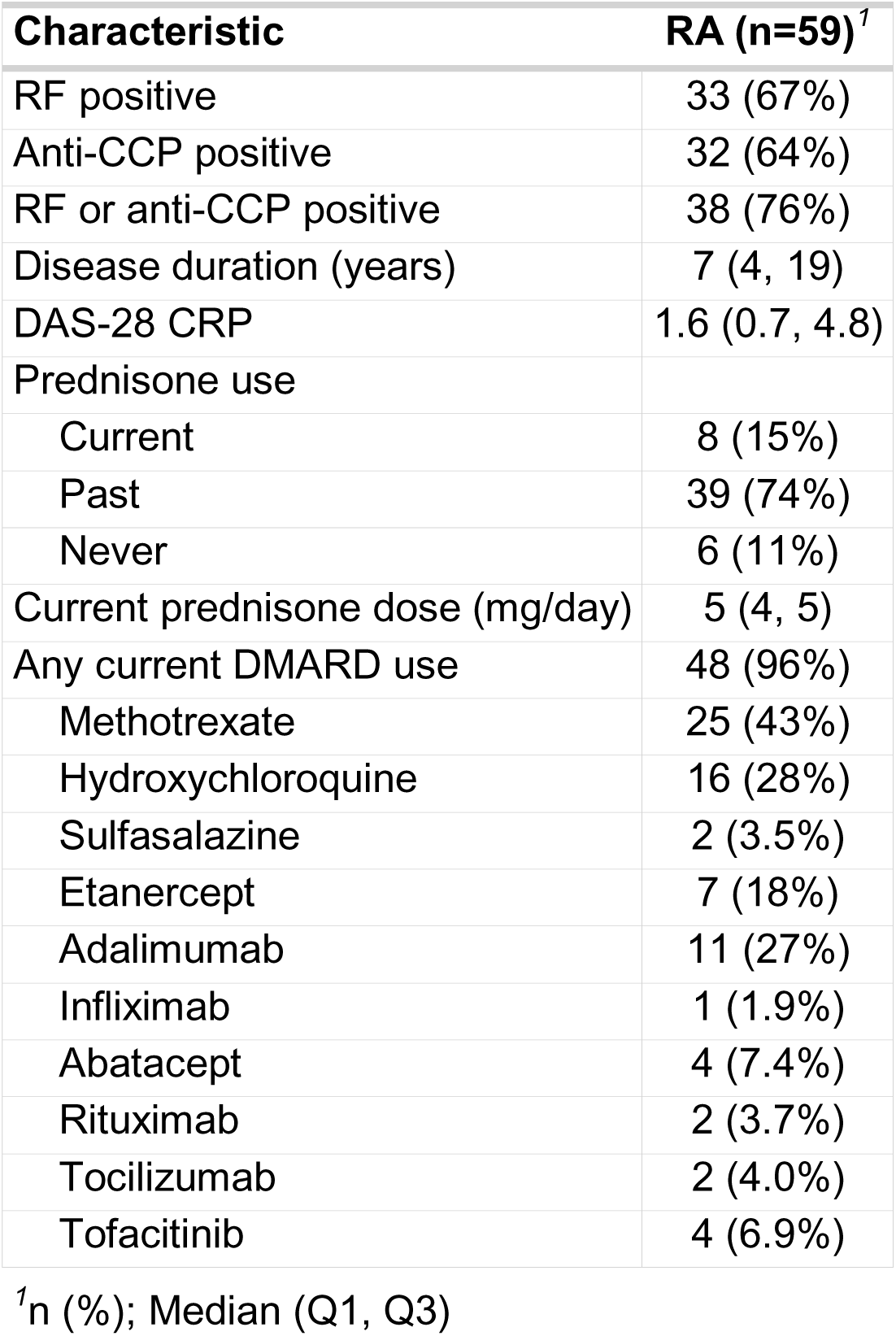
Clinical Characteristics of Participants with RA (n=59)

### Plasma Fatty Acid Profiles

Plasma MUFAs (Figure 1A) were elevated in RA patients, including palmitoleic acid [1.93% (IQR 1.36–2.74) vs 1.46% (IQR 1.17–2.14), p = 0.022] and oleic acid [18.73% (IQR 17.18–20.42) vs 16.87% (IQR 14.91–18.19), p = 0.001], alongside higher SCD-1 index [2.67 (IQR 2.38–2.93) vs 2.21 (IQR 1.93–2.79), p = 0.010], suggesting a pattern consistent with enhanced endogenous MUFA biosynthesis in these patients. In contrast, n-6 PUFAs (Figure 1B), including linoleic acid [31.59% (IQR 28.89–33.33) vs 33.03% (IQR 30.86–35.67), p = 0.035] and arachidonic acid [7.28% (IQR 6.43–8.21) vs 8.51% (IQR 7.19–9.67), p = 0.004], were lower in RA, potentially reflecting preferential utilization in pro-inflammatory or oxidative pathways. No significant differences were observed in saturated fatty acids (Figure 1C) or n-3 PUFAs (Figure 1D), suggesting that changes in fatty acid composition may be limited to specific lipid classes.

**Figure 1.**
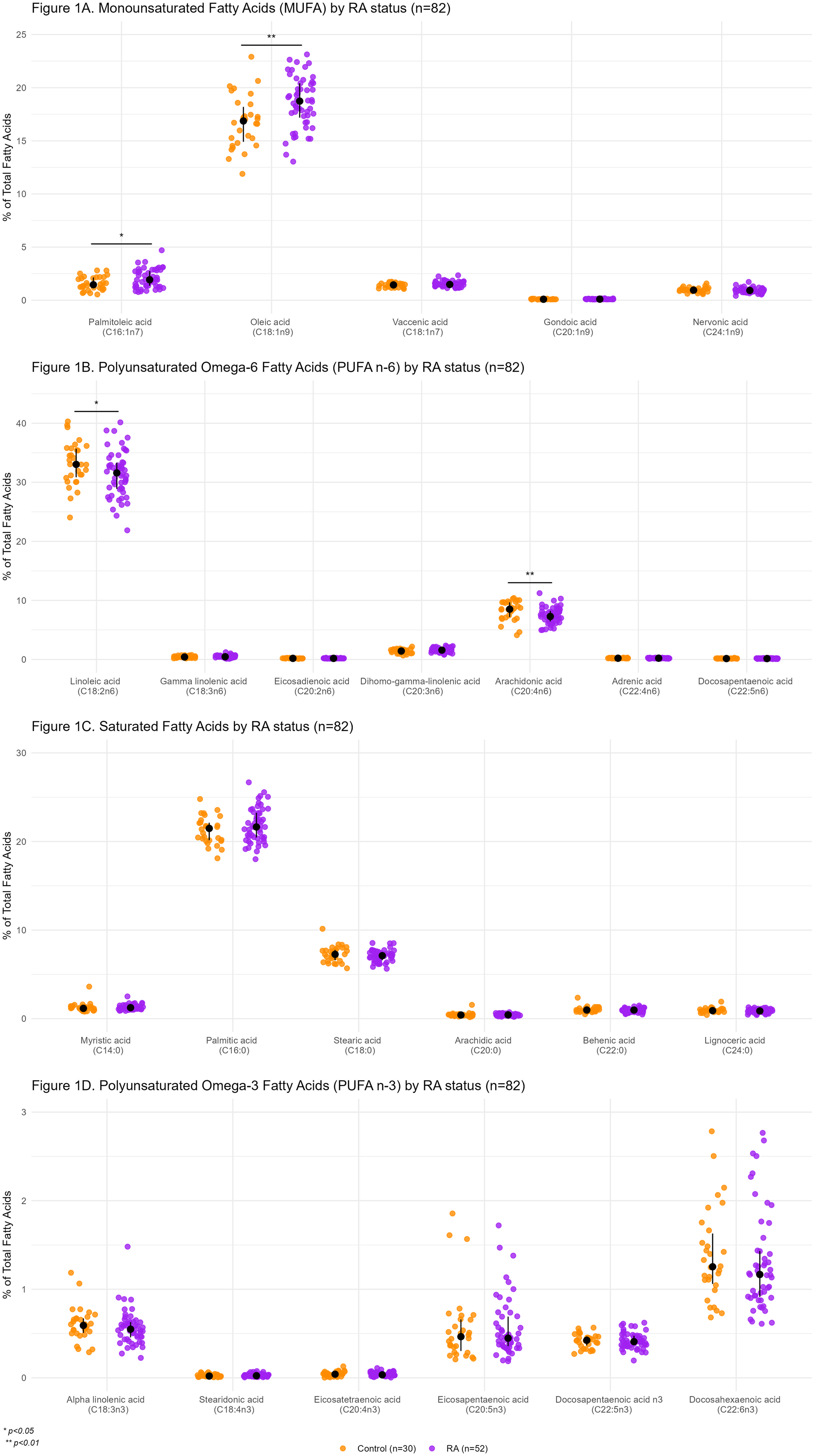
Summary of Fatty Acids Concentration by RA Status (n=82)

### Dietary Intake

Overall diet quality, assessed by the energy-adjusted HEI-2015 score, was comparable between individuals with RA and controls (p = 0.7) (Table 1). There were no significant differences in average dietary intake (Supplemental Table 1) of palmitoleic, oleic, gondoic, linoleic, or arachidonic acids between RA patients and controls (all p > 0.5). These findings indicate that plasma fatty acid differences cannot be attributed solely to differences in dietary intake and are consistent with disease-related metabolic alterations.

### Correlations with Clinical Parameters

Spearman correlation analyses (Supplemental Table 2) indicated that plasma MUFAs (palmitoleic, oleic, and gondoic acids) were positively associated with RA disease activity (DAS28-CRP) and measures of adiposity, including BMI, FMI, and waist-to-hip ratio, and inversely associated with insulin sensitivity. Linoleic acid was inversely associated with waist-to-hip ratio and positively correlated with insulin sensitivity, supporting a potentially protective role. Arachidonic acid did not show significant associations with disease activity, adiposity, or metabolic markers in either RA or controls, indicating no clear relationship between circulating AA levels and these clinical measures in this cohort.

### Multivariable Analyses

In multivariable models, higher plasma oleic (Table 3) and gondoic acids (Table 4) were positively associated with DAS28-CRP independent of dietary intake. Each 1% increase in oleic acid corresponded to a 0.65-point higher DAS28-CRP (95% CI 0.11–1.20, p = 0.019), and each 1% increase in gondoic acid to a 0.0108-point higher DAS28-CRP (95% CI 0.0043–0.0173, p = 0.002). Higher plasma oleic acid was also linked to adiposity, with a 0.01-unit increase in waist-to-hip ratio associated with a 13% higher oleic acid level (95% CI 4.5–21, p = 0.003).

**Table 3.**
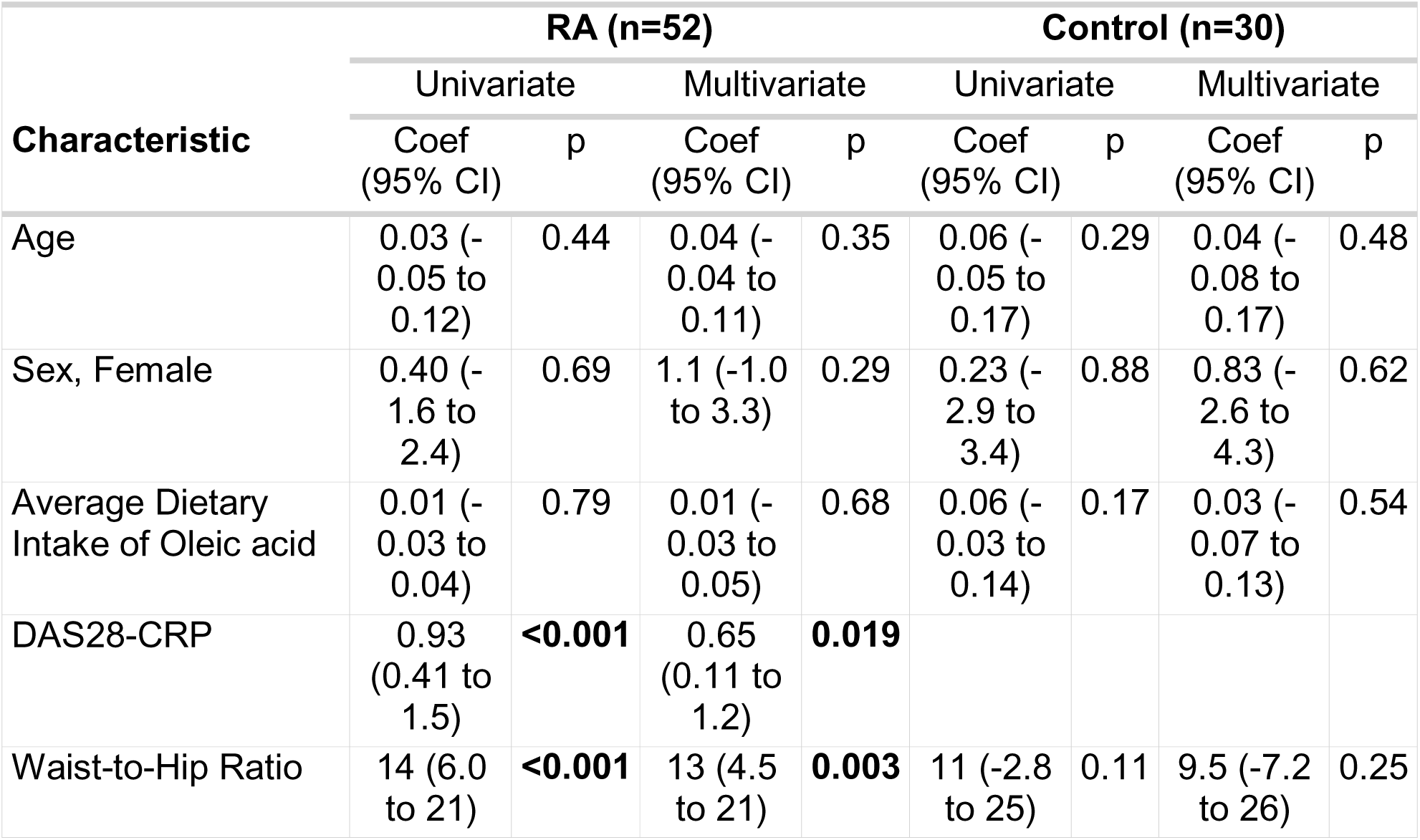
Summary of Associations between Oleic acid (C18:1n9) and Age, Sex, Average Dietary Intake of Oleic acid, Waist-to-hip Ratio and DAS28-CRP by RA Status (n=82)

**Table 4.**
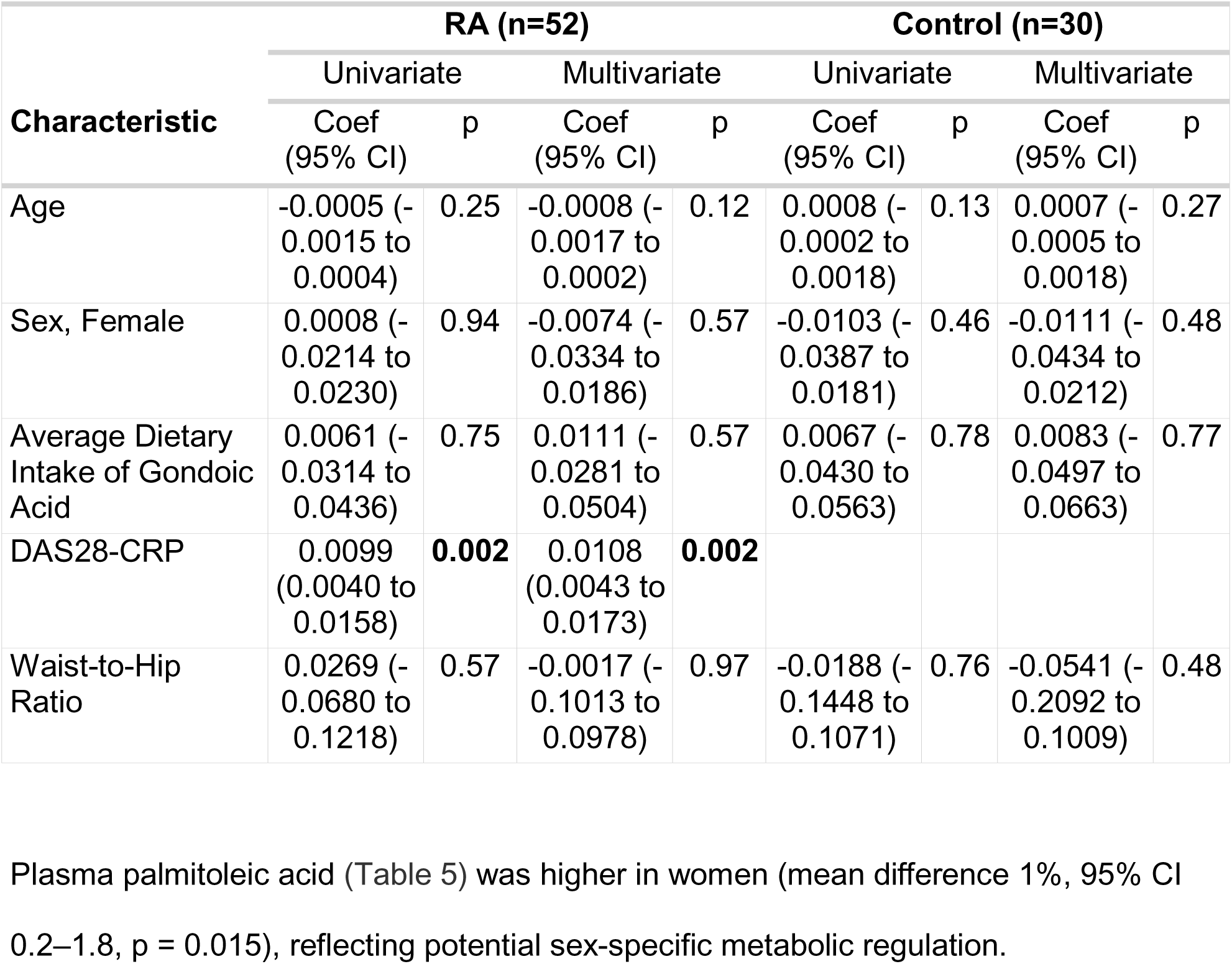
Summary of Associations between Gondoic Acid (C20:1n9) and Age, Sex, Average Dietary Intake of Gondoic Acid, Waist-to-hip Ratio and DAS28-CRP by RA Status (n=82)

**Table 5.**
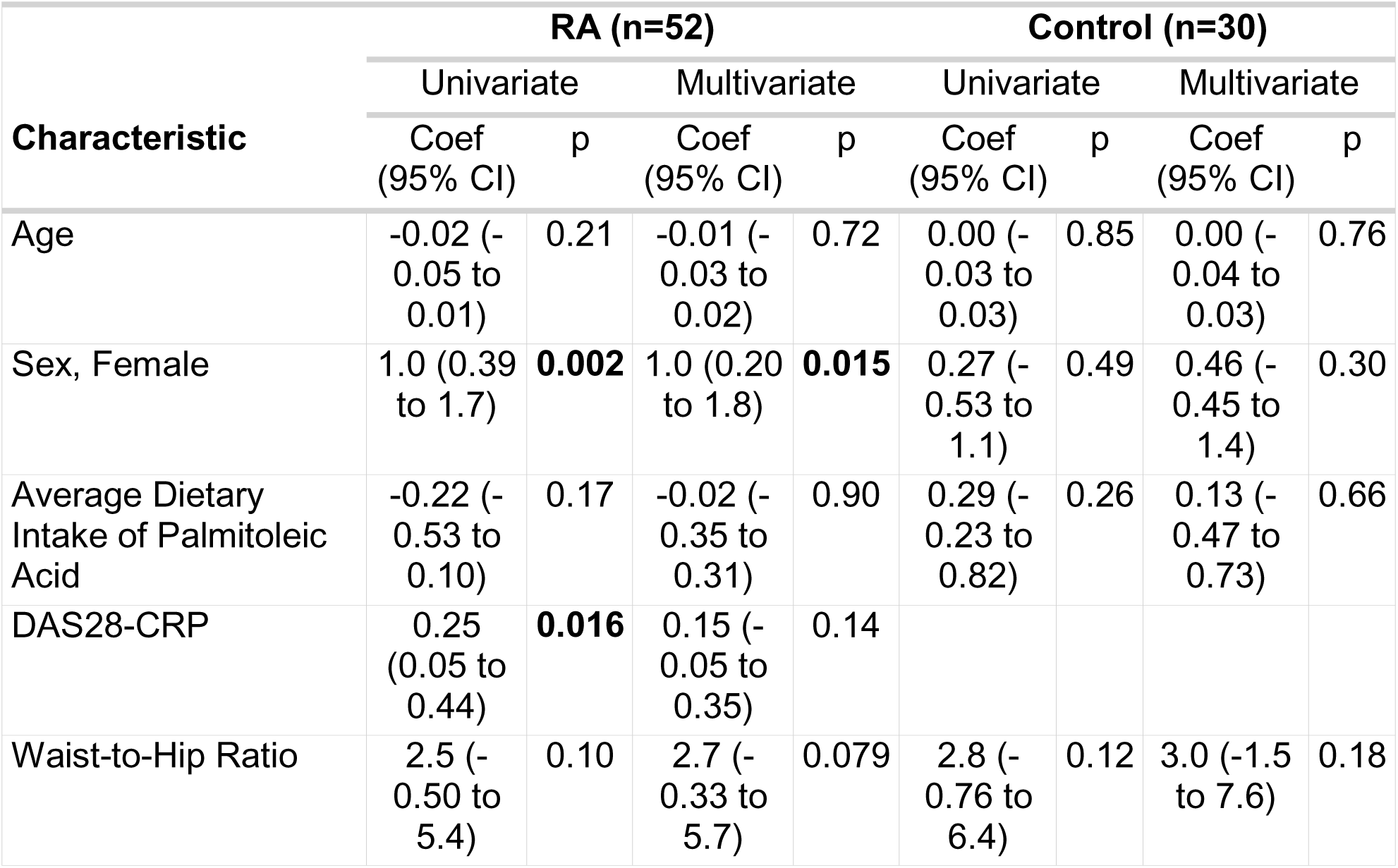
Summary of Associations between Plasma Palmitoleic Acid (C16:1n7) and Age, Sex, Dietary Intake of Palmitoleic Acid, Waist-to-hip Ratio and DAS28-CRP by RA Status (n=82) Higher linoleic acid (Table 6) was negatively associated with waist-to-hip ratio in both RA (−15.332; 95% CI –29.727 to –0.9363, p = 0.037) and controls (−37.204; 95% CI – 57.237 to –17.171, p < 0.001), whereas plasma arachidonic acid (Table 7) was not significantly associated with disease activity or central adiposity. Collectively, these results highlight a disease-specific fatty acid metabolic signature in RA, characterized by elevated MUFAs linked to both adiposity and inflammation, while certain PUFAs may confer metabolic benefits.

**Table 6.**
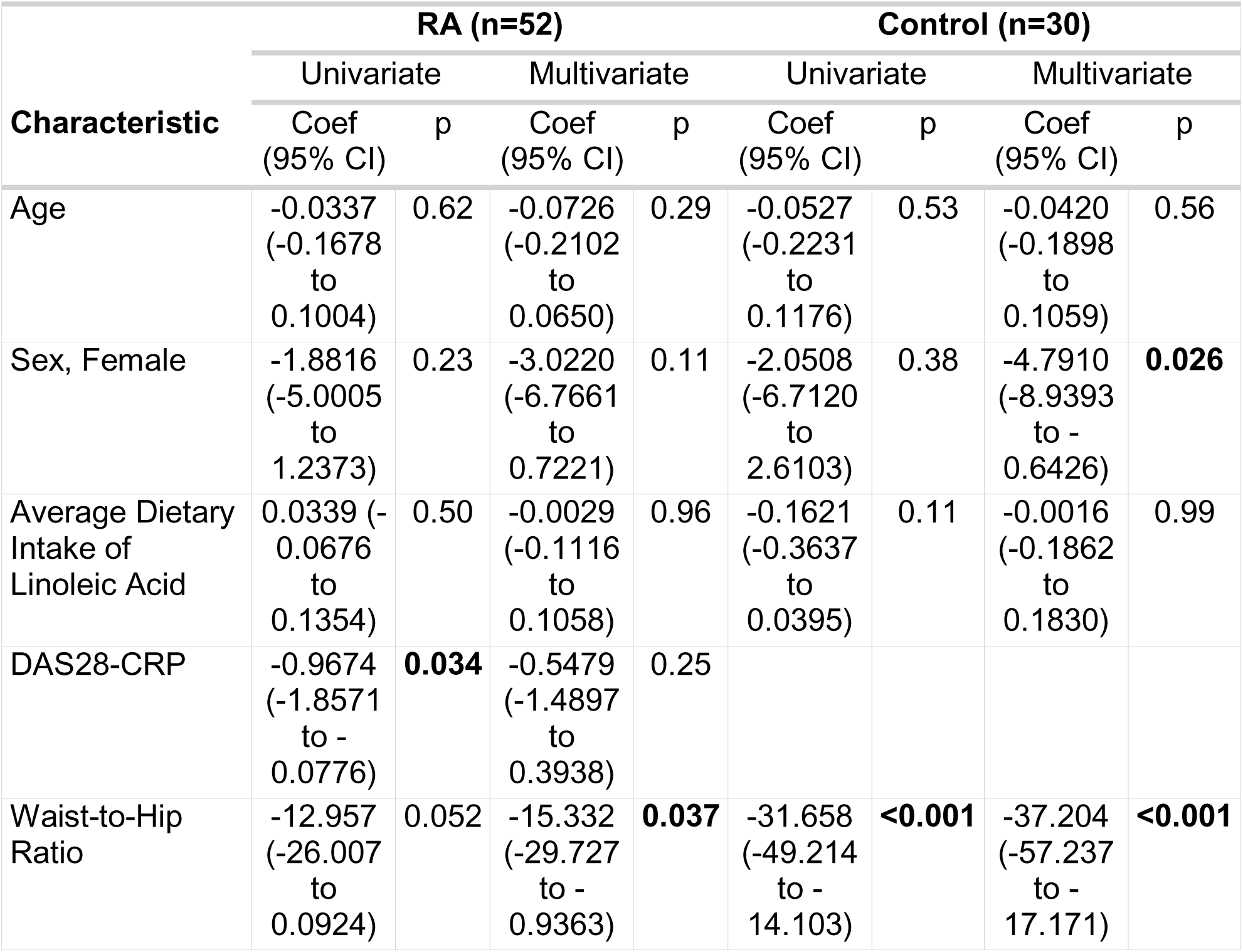
Summary of Associations between Linoleic Acid (C18:2n6) and Age, Sex, Average Dietary Intake of Linoleic Acid, Waist-to-hip Ratio and DAS28-CRP by RA Status (n=82)

**Table 7.**
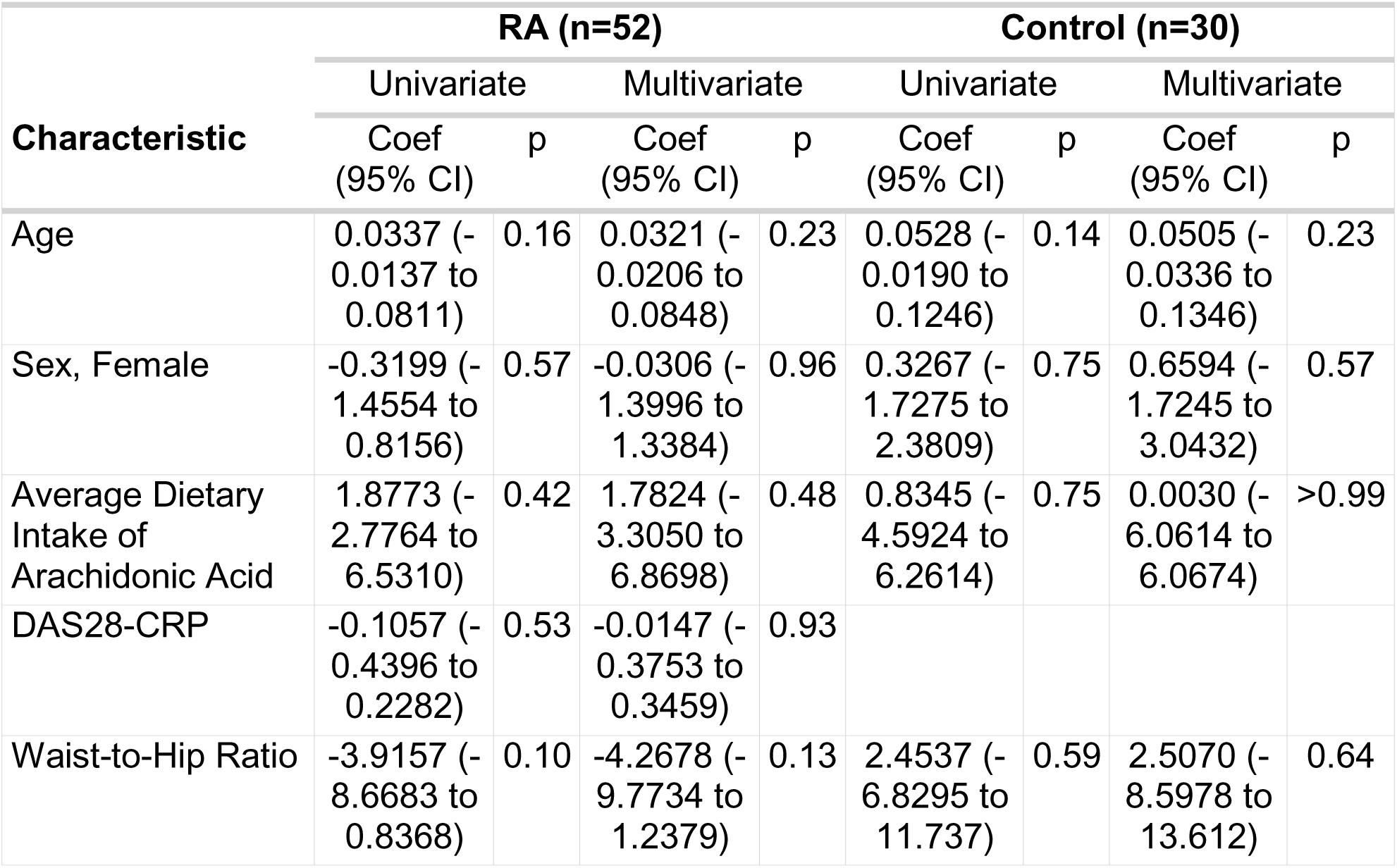
Summary of Associations between Plasma Arachidonic Acid (C20:4n6) and Age, Sex, Dietary Intake of Arachidonic Acid, Waist-to-hip Ratio and DAS28-CRP by RA Status (n=82)

## Discussion

This study demonstrates significant alterations in plasma MUFAs in RA, with oleic and gondoic acid levels positively associated with disease activity. These associations persisted after adjusting for dietary intake and overall diet quality, suggesting that non-dietary factors, including endogenous metabolic regulation, may contribute to the observed lipid differences, although causality cannot be inferred from this cross-sectional analysis.

Elevated SCD-1 index in RA patients, reflected by the oleic-to-stearic acid ratio, indicates increased endogenous MUFA synthesis. SCD-1 converts saturated fatty acids — palmitic and stearic acids — into MUFAs oleic and palmitoleic acids, and has been linked to metabolic and inflammatory conditions, including obesity and insulin resistance. In RA, enhanced SCD-1 index may support the metabolic demands of activated immune cells and influence lipid signaling pathways, including macrophage polarization and cytokine production.

SCD-1 is highly expressed in the liver and adipose tissues, which contribute most to circulating MUFA levels (26). It is also present in immune cells, including macrophages, T cells, dendritic cells, and B cells, where expression increases during activation and proliferation to support membrane remodeling, energy metabolism, and signaling (27–29). Through its role in regulating systemic and cell-specific MUFA synthesis, SCD-1 is positioned at the interface of metabolic regulation and immune function, offering a biologically plausible link between lipid metabolism and inflammatory pathways relevant to RA.

Sex-specific differences were observed, with higher palmitoleic acid levels in women with RA. This may reflect interactions between estrogen, adiposity, and SCD-1–mediated lipogenesis. While estrogen has been reported to repress SCD-1 in some contexts, inflammatory and metabolic alterations in RA, along with higher fat mass in women, may provide substrates or signaling cues that override this repression, facilitating increased palmitoleic acid synthesis(30). This sex difference was absent in controls, highlighting a disease-specific interplay between hormonal and metabolic regulation.

RA participants also exhibited reduced plasma n-6 PUFAs, including linoleic and arachidonic acids, likely reflecting preferential utilization in pro-inflammatory pathways (e.g., eicosanoid biosynthesis) and oxidative degradation under chronic inflammation. In contrast, n-3 PUFAs and saturated fatty acids did not differ between RA and controls, suggesting a selective depletion of n-6 PUFAs rather than a global change in fatty acid composition. A lower status of n-6 fatty acids has been observed in populations at risk for RA, where higher erythrocyte levels of linoleic acid were associated with a lower incidence of subsequent RA, indicating disease-related effects on n-6 PUFA metabolism (31). Studies of fatty acid metabolism also highlight the central role of arachidonic acid–derived eicosanoids in RA inflammation, providing biological support for the idea that chronic systemic inflammation may preferentially impact n-6 PUFA pools(32). Together, these findings suggest a shift in lipid metabolism toward pro-inflammatory and catabolic pathways, consistent with sustained systemic inflammation in RA.

Interestingly, higher circulating MUFA levels were associated with greater RA disease activity. While dietary MUFAs are generally considered protective, elevated endogenously produced MUFAs, driven by SCD-1 activity, may not confer the same benefits. In RA, increased MUFA levels may reflect compensatory SCD-1–mediated synthesis in response to reduced n-6 PUFA availability or altered metabolic status, including increased abdominal adiposity.

Obesity is common among patients with RA and is linked to worse disease outcomes, including higher disease activity and greater functional disability(33, 34). Chronic inflammatory signaling, altered energy demands, and adiposity-related hormones (e.g., leptin) may further modulate SCD-1 activity, contributing to dysregulated MUFA metabolism(26, 35, 36). RA participants also exhibited a less favorable metabolic profile, including elevated triglycerides, a trend toward lower insulin sensitivity, and lower physical activity, which may exacerbate alterations in MUFA metabolism and higher disease activity.

Given the central role of SCD-1 in MUFA synthesis and its association with RA disease activity, pharmacologic inhibition of SCD-1 represents a potential therapeutic avenue(36). While SCD-1 inhibitors are currently under investigation in preclinical and early clinical studies for metabolic and inflammatory conditions, further research is needed to evaluate their safety, efficacy, and immunomodulatory effects in RA(37, 38).

In controls, plasma palmitoleic, oleic, and gondoic acid levels were not associated with age, sex, dietary intake, or waist-to-hip ratio, supporting the disease-specific nature of these associations. However, the smaller control sample may have limited the power to detect subtle associations.

Our observation that higher plasma linoleic acid (% of total fatty acids) was negatively associated with waist-to-hip ratio in both RA and control participants suggests that circulating linoleic acid may be linked to lower central adiposity, independent of disease status. This pattern is consistent with epidemiologic evidence showing that plasma linoleic acid is inversely associated with adiposity measures, including waist circumference and markers of metabolic dysregulation. For example, in a large cohort of Irish adults, plasma linoleic acid decreased with increasing adiposity and was inversely correlated with markers of inflammation and insulin resistance, indicating a healthier metabolic profile with higher linoleic acid levels (39). Furthermore, plasma and erythrocyte linoleic acid in other cohorts has been associated with lower central adiposity and reduced risk of features of metabolic syndrome(40). These reports are in keeping with our data suggesting that linoleic acid may reflect favorable adiposity and cardiometabolic status rather than simply dietary intake alone.

In contrast to the robust adiposity associations we observed for linoleic acid, plasma arachidonic acid was not significantly associated with DAS28-CRP or waist-to-hip ratio in either RA patients or controls. Arachidonic acid is a downstream n-6 PUFA and a well-known precursor for pro-inflammatory eicosanoids, and mechanistic studies have implicated its metabolites in inflammatory signaling pathways relevant to chronic inflammatory diseases such as RA. Eicosanoid pathways, which derive many bioactive mediators from arachidonic acid, are thought to play pivotal roles in the pathogenesis of rheumatic diseases (41). However, observational data relating circulating arachidonic acid directly to conventional inflammatory biomarkers in humans have been mixed.

Some population studies have found no association between serum arachidonic acid or other n-6 PUFAs and C-reactive protein levels, even though linoleic acid showed inverse associations with CRP(42). Other work has demonstrated that plasma PUFA profiles, including both n-3 and n-6 fatty acids, can be variably associated with pro– and anti-inflammatory markers depending on the context and covariates studied(43). Our null findings for arachidonic acid in relation to RA disease activity and central adiposity likely reflect this complexity, indicating that plasma arachidonic acid alone may not be a strong independent predictor of systemic inflammation or adiposity measures in these cohorts.

Limitations include the cross-sectional design, which precludes causal inference; plasma fatty acid measurements, reflecting short– to medium-term lipid status rather than long-term or tissue-specific metabolism; and potential unmeasured confounders such as medications, genetic variation, or dietary intake not captured accurately by self-report. Accordingly, differences in plasma fatty acids may reflect both disease-related metabolic alterations and limitations of dietary assessment. In addition, we did not measure markers of fatty acid oxidation or downstream inflammatory mediators in plasma, which could provide further insight into the functional consequences of altered fatty acid metabolism. This cohort was recruited from academic centers and consisted predominantly of women with established RA and a median BMI in the obese range.

Findings may not generalize to men, individuals with early untreated RA, or populations with different demographic or treatment distributions. Larger, longitudinal studies are needed to validate these findings and explore subgroup effects by sex, age, or treatment status.

Future directions should investigate tissue-specific roles of SCD-1 in immune cells and adipose tissue, and determine whether modulation of MUFA metabolism can influence inflammation or clinical outcomes. Integrating metabolic and immunologic profiling may provide new insights for personalized RA treatment strategies.

## Declarations

The study was conducted in accordance with the Declaration of Helsinki and approved by Institutional Review Board or Ethics Committee at University of Alabama at Birmingham, the Ohio State University and the University of Oklahoma Health Sciences Center (Approval numbers: F160426002, 2019H022 and 16539, respectively). All participants provided written informed consent prior to enrollment.

## Availability of data and materials

The datasets generated and/or analysed during the current study are not publicly available due to restrictions on participant privacy but are available from the corresponding author on reasonable request. Requests for access to the data should be directed to Beatriz Y. Hanaoka.

## Authors’ contributions

BH conceived and designed the study; oversaw participant recruitment; collected clinical data; performed physical assessments and disease activity scoring; supervised laboratory assays; interpreted the results; and drafted the manuscript. SNW performed laboratory assays, interpreted the results, and assisted with manuscript drafting. RMC conducted plasma fatty acid measurements and performed preliminary data analyses. MAB oversaw plasma fatty acid measurements and performed preliminary data analyses. KH collected dietary data, calculated the Healthy Eating Index (HEI), and interpreted dietary results. SP calculated stearoyl-CoA desaturase-1 index and performed statistical analyses. AK contributed to interpretation of the statistical analyses, including multiple regression modeling, and assisted with manuscript drafting. ZY and YMD assisted with interpretation of associations between fatty acids and clinical variables. All authors critically revised the manuscript for intellectual content and approved the final version.

## Data Availability

All data produced in the present study are available upon reasonable request to the authors.

## Acknowledgements

The authors would like to thank all study participants for their time and contribution to this research. We also acknowledge the support of the Clinical and Translational Science Institute at the Ohio State University and University of Alabama at Birmingham for providing facilities and technical assistance. The authors received no professional writing or editorial assistance beyond the use of a Large Language Model (ChatGPT, OpenAI, San Francisco, CA, USA) for language editing, which was supervised and verified by all authors.

## References

1. Giles JT, Allison M, Blumenthal RS, Post W, Gelber AC, Petri M, et al. Abdominal adiposity in rheumatoid arthritis: association with cardiometabolic risk factors and disease characteristics. Arthritis Rheum. 2010;62(11):3173–82.

2. McInnes IB, Schett G. The pathogenesis of rheumatoid arthritis. N Engl J Med. 2011;365(23):2205–19.

3. Calder PC. Omega-3 fatty acids and inflammatory processes: from molecules to man. Biochem Soc Trans. 2017;45(5):1105–15.

4. Calder PC, Grimble RF. Polyunsaturated fatty acids, inflammation and immunity. Eur J Clin Nutr. 2002;56 Suppl 3:S14–9.

5. Sigaux J, Mathieu S, Nguyen Y, Sanchez P, Letarouilly JG, Soubrier M, et al. Impact of type and dose of oral polyunsaturated fatty acid supplementation on disease activity in inflammatory rheumatic diseases: a systematic literature review and meta-analysis. Arthritis Res Ther. 2022;24(1):100.

6. Lourdudoss C, Di Giuseppe D, Wolk A, Westerlind H, Klareskog L, Alfredsson L, et al. Dietary Intake of Polyunsaturated Fatty Acids and Pain in Spite of Inflammatory Control Among Methotrexate-Treated Early Rheumatoid Arthritis Patients. Arthritis Care Res (Hoboken). 2018;70(2):205–12.

7. Veselinovic M, Vasiljevic D, Vucic V, Arsic A, Petrovic S, Tomic-Lucic A, et al. Clinical Benefits of n-3 PUFA and –Linolenic Acid in Patients with Rheumatoid Arthritis. Nutrients. 2017;9(4).

8. Geusens P, Wouters C, Nijs J, Jiang Y, Dequeker J. Long-term effect of omega-3 fatty acid supplementation in active rheumatoid arthritis. A 12-month, double-blind, controlled study. Arthritis Rheum. 1994;37(6):824–9.

9. Proudman SM, James MJ, Spargo LD, Metcalf RG, Sullivan TR, Rischmueller M, et al. Fish oil in recent onset rheumatoid arthritis: a randomised, double-blind controlled trial within algorithm-based drug use. Ann Rheum Dis. 2015;74(1):89–95.

10. Santa-Maria C, Lopez-Enriquez S, Montserrat-de la Paz S, Geniz I, Reyes-Quiroz ME, Moreno M, et al. Update on Anti-Inflammatory Molecular Mechanisms Induced by Oleic Acid. Nutrients. 2023;15(1).

11. Kris-Etherton PM. AHA science advisory: monounsaturated fatty acids and risk of cardiovascular disease. J Nutr. 1999;129(12):2280–4.

12. Mauvoisin D, Mounier C. Hormonal and nutritional regulation of SCD1 gene expression. Biochimie. 2011;93(1):78–86.

13. Attie AD, Krauss RM, Gray-Keller MP, Brownlie A, Miyazaki M, Kastelein JJ, et al. Relationship between stearoyl-CoA desaturase activity and plasma triglycerides in human and mouse hypertriglyceridemia. J Lipid Res. 2002;43(11):1899–907.

14. Chow LS, Li S, Eberly LE, Seaquist ER, Eckfeldt JH, Hoogeveen RC, et al. Estimated plasma stearoyl co-A desaturase-1 activity and risk of incident diabetes: the Atherosclerosis Risk in Communities (ARIC) study. Metabolism. 2013;62(1):100–8.

15. Waters KM, Ntambi JM. Polyunsaturated fatty acids inhibit hepatic stearoyl-CoA desaturase-1 gene in diabetic mice. Lipids. 1996;31 Suppl:S33–6.

16. Waters KM, Ntambi JM. Insulin and dietary fructose induce stearoyl-CoA desaturase 1 gene expression of diabetic mice. J Biol Chem. 1994;269(44):27773–7.

17. Aletaha D, Neogi T, Silman AJ, Funovits J, Felson DT, Bingham CO, 3rd, et al. 2010 rheumatoid arthritis classification criteria: an American College of Rheumatology/European League Against Rheumatism collaborative initiative. Ann Rheum Dis. 2010;69(9):1580–8.

18. Matsuda M, DeFronzo RA. Insulin sensitivity indices obtained from oral glucose tolerance testing: comparison with the euglycemic insulin clamp. Diabetes Care. 1999;22(9):1462–70.

19. (CDC) CfDC. Centers for Disease Control and Prevention (CDC), National Center for Health Statistics. National Health and Nutrition Examination Survey III: Body Measurements (Anthropometry) Manual. Hyattsville, MD: U.S. Department of Health and Human Services; 1988-1994.

20. Folch J, Lees M, Sloane Stanley GH. A simple method for the isolation and purification of total lipides from animal tissues. J Biol Chem. 1957;226(1):497–509.

21. Stoffel W, Chu, F., and Ahrens, E.H. Jr. Analysis of long-chain fatty acids by gas-liquid chromatography. Analytical Chemistry. 1959;31(2):307–8.

22. Arnold LE, Young AS, Belury MA, Cole RM, Gracious B, Seidenfeld AM, et al. Omega-3 Fatty Acid Plasma Levels Before and After Supplementation: Correlations with Mood and Clinical Outcomes in the Omega-3 and Therapy Studies. J Child Adolesc Psychopharmacol. 2017;27(3):223–33.

23. Craig CL, Marshall AL, Sjostrom M, Bauman AE, Booth ML, Ainsworth BE, et al. International physical activity questionnaire: 12-country reliability and validity. Med Sci Sports Exerc. 2003;35(8):1381–95.

24. Schakel SF, Sievert YA, Buzzard IM. Sources of data for developing and maintaining a nutrient database. J Am Diet Assoc. 1988;88(10):1268–71.

25. Krebs-Smith SM, Pannucci TE, Subar AF, Kirkpatrick SI, Lerman JL, Tooze JA, et al. Update of the Healthy Eating Index: HEI-2015. J Acad Nutr Diet. 2018;118(9):1591–602.

26. Liu X, Strable MS, Ntambi JM. Stearoyl CoA desaturase 1: role in cellular inflammation and stress. Adv Nutr. 2011;2(1):15–22.

27. Bogie JFJ, Grajchen E, Wouters E, Corrales AG, Dierckx T, Vanherle S, et al. Stearoyl-CoA desaturase-1 impairs the reparative properties of macrophages and microglia in the brain. J Exp Med. 2020;217(5).

28. Grajchen E, Loix M, Baeten P, Corte-Real BF, Hamad I, Vanherle S, et al. Fatty acid desaturation by stearoyl-CoA desaturase-1 controls regulatory T cell differentiation and autoimmunity. Cell Mol Immunol. 2023;20(6):666–79.

29. Zhou X, Zhu X, Li C, Li Y, Ye Z, Shapiro VS, et al. Stearoyl-CoA Desaturase-Mediated Monounsaturated Fatty Acid Availability Supports Humoral Immunity. Cell Rep. 2021;34(1):108601.

30. Belkaid A, Duguay SR, Ouellette RJ, Surette ME. 17beta-estradiol induces stearoyl-CoA desaturase-1 expression in estrogen receptor-positive breast cancer cells. BMC Cancer. 2015;15:440.

31. de Pablo P, Romaguera D, Fisk HL, Calder PC, Quirke AM, Cartwright AJ, et al. High erythrocyte levels of the n-6 polyunsaturated fatty acid linoleic acid are associated with lower risk of subsequent rheumatoid arthritis in a southern European nested case-control study. Ann Rheum Dis. 2018;77(7):981–7.

32. Calder PC. Polyunsaturated fatty acids and inflammatory processes: New twists in an old tale. Biochimie. 2009;91(6):791–5.

33. Baker JF, England BR, Mikuls TR, Sayles H, Cannon GW, Sauer BC, et al. Obesity, Weight Loss, and Progression of Disability in Rheumatoid Arthritis. Arthritis Care Res (Hoboken). 2018;70(12):1740–7.

34. Gremese E, Carletto A, Padovan M, Atzeni F, Raffeiner B, Giardina AR, et al. Obesity and reduction of the response rate to anti-tumor necrosis factor alpha in rheumatoid arthritis: an approach to a personalized medicine. Arthritis Care Res (Hoboken). 2013;65(1):94–100.

35. Biddinger SB, Miyazaki M, Boucher J, Ntambi JM, Kahn CR. Leptin suppresses stearoyl-CoA desaturase 1 by mechanisms independent of insulin and sterol regulatory element-binding protein-1c. Diabetes. 2006;55(7):2032–41.

36. Sun Q, Xing X, Wang H, Wan K, Fan R, Liu C, et al. SCD1 is the critical signaling hub to mediate metabolic diseases: Mechanism and the development of its inhibitors. Biomed Pharmacother. 2024;170:115586.

37. Naghashi N, Babaei E, Sanaat Z, Mehdizadeh A. Investigating the effect of stearoyl-coenzyme desaturase 1 Inhibition on inflammatory and anti-inflammatory patterns in luminal A breast cancer patients’ peripheral blood mononuclear cells. Discov Oncol. 2025;16(1):2095.

38. Kirad S, Puri S, Deepa PR, Sankaranarayanan M. An insight into advances and challenges in the development of potential stearoyl Co-A desaturase 1 inhibitors. RSC Adv. 2024;14(41):30487–517.

39. Li K, Brennan L, Bloomfield JF, Duff DJ, McNulty BA, Flynn A, et al. Adiposity Associated Plasma Linoleic Acid is Related to Demographic, Metabolic Health and Haplotypes of FADS1/2 Genes in Irish Adults. Mol Nutr Food Res. 2018;62(7):e1700785.

40. Petersen KS, Sullivan VK, Fulgoni VL, 3rd, Eren F, Cassens ME, Bunczek MT, et al. Circulating Concentrations of Essential Fatty Acids, Linoleic and alpha-Linolenic Acid, in US Adults in 2003-2004 and 2011-2012 and the Relation with Risk Factors for Cardiometabolic Disease: An NHANES Analysis. Curr Dev Nutr. 2020;4(10):nzaa149.

41. Korotkova M, Jakobsson PJ. Persisting eicosanoid pathways in rheumatic diseases. Nat Rev Rheumatol. 2014;10(4):229–41.

42. Virtanen JK, Mursu J, Voutilainen S, Tuomainen TP. The associations of serum n-6 polyunsaturated fatty acids with serum C-reactive protein in men: the Kuopio Ischaemic Heart Disease Risk Factor Study. Eur J Clin Nutr. 2018;72(3):342–8.

43. Ferrucci L, Cherubini A, Bandinelli S, Bartali B, Corsi A, Lauretani F, et al. Relationship of plasma polyunsaturated fatty acids to circulating inflammatory markers. J Clin Endocrinol Metab. 2006;91(2):439–46.

