## Supplemental Tables 1 and 2 for "Elevated Plasma Monounsaturated Fatty Acids and Their Associations with Disease Activity, Adiposity, and Sex in Patients with Rheumatoid Arthritis: A Cross-Sectional Study"

**Supplemental Table 1.** Average Dietary Intake of Fatty Acids (n=92)

| **Average Dietary Intake** | **Control (n=33)** | **RA (n=59)** | **p** |
| --- | --- | --- | --- |
| Average Dietary Intake of Palmitoleic acid (C16:1n7) | 0.10 (0.06, 1.16) | 0.10 (0.06, 0.97) | 0.7 |
| Average Dietary Intake of Oleic acid (C18:1n9) | 3 (1, 20) | 2 (1, 21) | >0.9 |
| Average Dietary Intake of Gondoic acid (C20:1n9) | 0.03 (0.02, 0.24) | 0.03 (0.02, 0.21) | 0.9 |
| Average Dietary Intake of Linoleic acid (C18:2n6) | 2 (1, 12) | 2 (1, 13) | >0.9 |
| Average Dietary Intake of Arachidonic acid (C20:4n6) | 0.01 (0.01, 0.15) | 0.01 (0.01, 0.12) | 0.5 |

**Supplemental Table 2.** Correlations between Fatty Acids and Clinical Parameters among RA and Control Patients (n=82)

| **Fatty Acids** | **Parameters** | **Correlation Coef** | **p** | **Correlation Coef** | **p** |
| --- | --- | --- | --- | --- | --- |
| Palmitoleic acid (C16:1n7) | Age | -0.176 | 0.211 | -0.037 | 0.847 |
| Palmitoleic acid (C16:1n7) | BMI | 0.356 | 0.010 | 0.224 | 0.235 |
| Palmitoleic acid (C16:1n7) | CRP | 0.264 | 0.067 | 0.592 | 0.001 |
| Palmitoleic acid (C16:1n7) | DAS28-CRP | 0.334 | 0.016 |  | NA |
| Palmitoleic acid (C16:1n7) | FFMI | 0.093 | 0.514 | 0.157 | 0.415 |
| Palmitoleic acid (C16:1n7) | FMI | 0.434 | 0.001 | 0.402 | 0.031 |
| Palmitoleic acid (C16:1n7) | HbA1c | 0.053 | 0.723 | 0.167 | 0.395 |
| Palmitoleic acid (C16:1n7) | Matsuda Index | -0.400 | 0.003 | -0.253 | 0.177 |
| Palmitoleic acid (C16:1n7) | Energy adjusted HEI 2015 score | -0.353 | 0.012 | -0.276 | 0.155 |
| Palmitoleic acid (C16:1n7) | Total METS | -0.151 | 0.286 | -0.207 | 0.273 |
| Palmitoleic acid (C16:1n7) | Waist-to-hip ratio | 0.230 | 0.102 | 0.297 | 0.118 |
| Oleic acid (C18:1n9) | Age | 0.108 | 0.444 | 0.198 | 0.294 |
| Oleic acid (C18:1n9) | BMI | 0.075 | 0.596 | 0.134 | 0.479 |
| Oleic acid (C18:1n9) | CRP | 0.298 | 0.037 | 0.134 | 0.480 |
| Oleic acid (C18:1n9) | DAS28-CRP | 0.450 | 0.001 |  | NA |
| Oleic acid (C18:1n9) | FFMI | 0.060 | 0.673 | 0.192 | 0.319 |
| Oleic acid (C18:1n9) | FMI | 0.076 | 0.597 | 0.140 | 0.470 |
| Oleic acid (C18:1n9) | HbA1c | -0.195 | 0.190 | 0.137 | 0.488 |
| Oleic acid (C18:1n9) | Matsuda Index | -0.339 | 0.014 | -0.051 | 0.790 |
| Oleic acid (C18:1n9) | Energy adjusted HEI 2015 score | -0.172 | 0.233 | 0.158 | 0.423 |
| Oleic acid (C18:1n9) | Total METS | 0.050 | 0.725 | -0.196 | 0.300 |
| Oleic acid (C18:1n9) | Waist-to-hip ratio | 0.454 | 0.001 | 0.301 | 0.113 |
| Linoleic acid (C18:2n6) | Age | -0.071 | 0.616 | -0.119 | 0.531 |
| Linoleic acid (C18:2n6) | BMI | -0.144 | 0.309 | -0.290 | 0.120 |
| Linoleic acid (C18:2n6) | CRP | -0.255 | 0.077 | -0.333 | 0.073 |
| Linoleic acid (C18:2n6) | DAS28-CRP | -0.295 | 0.034 |  | NA |
| Linoleic acid (C18:2n6) | FFMI | -0.061 | 0.673 | -0.211 | 0.272 |
| Linoleic acid (C18:2n6) | FMI | -0.158 | 0.269 | -0.169 | 0.379 |
| Linoleic acid (C18:2n6) | HbA1c | 0.004 | 0.980 | -0.153 | 0.438 |
| Linoleic acid (C18:2n6) | Matsuda Index | 0.455 | 0.001 | 0.181 | 0.338 |
| Linoleic acid (C18:2n6) | Energy adjusted HEI 2015 score | 0.219 | 0.127 | -0.085 | 0.667 |
| Linoleic acid (C18:2n6) | Total METS | 0.081 | 0.566 | -0.014 | 0.942 |
| Linoleic acid (C18:2n6) | Waist-to-hip ratio | -0.271 | 0.052 | -0.580 | 0.001 |
| Gondoic acid (C20:1n9) | Age | -0.164 | 0.246 | 0.285 | 0.127 |
| Gondoic acid (C20:1n9) | BMI | -0.201 | 0.154 | -0.076 | 0.691 |
| Gondoic acid (C20:1n9) | CRP | 0.031 | 0.834 | -0.221 | 0.241 |
| Gondoic acid (C20:1n9) | DAS28-CRP | 0.428 | 0.002 |  | NA |
| Gondoic acid (C20:1n9) | FFMI | -0.158 | 0.268 | -0.101 | 0.601 |
| Gondoic acid (C20:1n9) | FMI | -0.189 | 0.183 | -0.153 | 0.428 |
| Gondoic acid (C20:1n9) | HbA1c | -0.412 | 0.004 | -0.002 | 0.990 |
| Gondoic acid (C20:1n9) | Matsuda Index | -0.061 | 0.669 | 0.338 | 0.068 |
| Gondoic acid (C20:1n9) | Energy adjusted HEI 2015 score | 0.130 | 0.369 | 0.421 | 0.026 |
| Gondoic acid (C20:1n9) | Total METS | -0.066 | 0.640 | -0.506 | 0.004 |
| Gondoic acid (C20:1n9) | Waist-to-hip ratio | 0.080 | 0.571 | -0.059 | 0.761 |
| Arachidonic acid (C20:4n6) | Age | 0.198 | 0.160 | 0.274 | 0.143 |
| Arachidonic acid (C20:4n6) | BMI | -0.248 | 0.076 | 0.029 | 0.880 |
| Arachidonic acid (C20:4n6) | CRP | -0.265 | 0.066 | -0.231 | 0.219 |
| Arachidonic acid (C20:4n6) | DAS28-CRP | -0.090 | 0.528 |  | NA |
| Arachidonic acid (C20:4n6) | FFMI | -0.235 | 0.097 | -0.151 | 0.434 |
| Arachidonic acid (C20:4n6) | FMI | -0.242 | 0.088 | -0.095 | 0.624 |
| Arachidonic acid (C20:4n6) | HbA1c | 0.183 | 0.217 | 0.143 | 0.468 |
| Arachidonic acid (C20:4n6) | Matsuda Index | 0.048 | 0.737 | 0.017 | 0.928 |
| Arachidonic acid (C20:4n6) | Energy adjusted HEI 2015 score | 0.145 | 0.315 | 0.164 | 0.404 |
| Arachidonic acid (C20:4n6) | Total METS | -0.123 | 0.386 | -0.059 | 0.758 |
| Arachidonic acid (C20:4n6) | Waist-to-hip ratio | -0.228 | 0.104 | 0.104 | 0.592 |
